# Drivers of antimicrobial prescriptions in hospitals from Asian low, middle and high income countries and implications for antibiotic stewardship

**DOI:** 10.64898/2026.04.07.26350373

**Authors:** Srishti Chhabra, Smeetha Nair, Alexandra Bramley, Chee Jia Yi, Kithalakshmi Vignesvaran, Daniel Rui En See, Louisa Jin Sun, Ann Hui Ching, Andrew Yunkai Li, Gyan Kayastha, Ploenchan Chetchotisakd, Ben S Cooper, Esmita Charani, Yin Mo

## Abstract

**Background:** Antibiotic use is prevalent in hospitals, driving the emergence of drug-resistant pathogens. We investigated the contextual influences on antibiotic prescribing behaviour across hospitals in high, middle, and low-income countries in Asia with an aim to provide actionable insights to improve prescribing behaviour.

**Methods:** We conducted a large qualitative study across ten institutions in Singapore, Nepal, and Thailand. Semi-structured interviews and ethnographic observations involving physicians, nurses, pharmacists, and management staff were conducted. Data were analysed thematically using QSR NVivo 14.

**Findings:** A total of 194 interviews were conducted amongst physicians (54·1%), nurses (19·6%), pharmacists (12·4%), and management staff (13·9%). Structural factors such as limited microbiology laboratory capabilities, concerns about antibiotic quality, weak infection prevention and control policies, and the lack of relevant, updated guidelines were prominent drivers for prolonged and broad-spectrum antibiotics prescriptions. Where these system supports were in place, prescribing decisions were less defensive and more targeted, although prescriber responsibility and concerns about immediate patient deterioration continued to influence practice. Across settings, clinicians tended to prioritise short-term perceived benefits of antibiotic treatment over the longer-term risks of antimicrobial resistance.

**Interpretation:** Efforts to optimise antibiotic prescribing should move beyond individual behaviour change and address system conditions that shape clinical decision-making. Strengthening diagnostics, infection prevention and control, antibiotic quality, and stewardship infrastructure can reduce prescriber uncertainty and enable safer, more context-specific antibiotic use.

**Funding:** The project was funded by Medical Research Council/ Department for International Development (MRC/DfID) (Grant Ref: MR/K006924/1) and Singapore National Medical Research Council (Grant Ref: CTGIIT18MAY-005)

## Introduction

Hospitals are critical hotspots for antimicrobial resistance (AMR)^1^. Approximately 35% of hospitalised patients receive at least one antibiotic during their stay, yet a significant portion of these prescriptions were deemed inappropriate^2^. The estimated five million AMR-related deaths in 2019, is projected to escalate to eight million deaths annually by 2050 without urgent action^3–6^, mainly driven by the elderly and severe multi-drug resistant hospital-acquired infections.

Antimicrobial stewardship programmes had been championed as a critical intervention to optimise prescribing, as highlighted by global initiatives such as the Jaipur Declaration and National Action Plan on AMR^7^. However, their success remains scarce, particularly in settings where systemic and financial barriers impede effective implementation and adoption^7–9^.

While individual physician factors, such as knowledge, prescribing experience, or patient-specific clinical presentations, are often targets for antibiotic prescribing behaviour changes, these alone fail to fully explain the observed disparities in prescribing practices across healthcare systems^10,11^. Prescribing decisions do not occur in isolation but are embedded within a broader ecosystem of institutional, cultural, and systemic influences. This underscores the need to reframe the issue of antibiotic prescribing not merely as a matter of individual physician behaviour but as one shaped by interdependent drivers within healthcare ecosystems. However, the interplay between these systemic influences and their cumulative impact on prescribing behaviour remains underexplored, particularly in resource-constrained environments^12^.

To bridge this gap, we conducted a multinational qualitative study to investigate the contextual and systemic influences on antibiotic prescribing behaviour across healthcare systems in high-, middle-, and low-income countries. By focusing on the healthcare ecosystem as a whole, this study aimed to illuminate the structural and cultural drivers that shape prescribing practices and provide generalisable and actionable insights to improve prescribing behaviour globally.

## Methods

### Study design

We conducted semi-structured interviews and ethnographic observations in 11 healthcare institutions across Singapore, Nepal, and Thailand. These institutions were at different stages along the spectrum of ASP implementation: mature, mid-implementation, and yet to implement, respectively. We enrolled adults aged 21 and above from five target groups, which included physicians (junior and senior), nurses, pharmacists and management staff.

### Sampling and recruitment of participants

We employed purposive sampling to recruit the initial participants for the study. To capture a wide range of perspectives, we applied snowballing as a secondary recruitment technique where each interviewee was asked to recommend additional participants. Participants were informed of their rights and provided audio recording consent prior to each interview.

### Data collection

Two separate semi-structured interview guides were created for healthcare providers and management staff. The interview guides were designed with insights from existing literature and domain experts from the three participating countries. Interviews were conducted face-to-face or over online conference calls. In Singapore, interviews were conducted in English; in Thailand and Nepal, they were conducted in local languages and later translated into English.

Ethnographic observations included ward rounds in both general wards and intensive care units (ICU), antibiotic stewardship meetings, physicians’ meetings and nursing handovers. No audio or video recordings were made during these observations. Detailed notes were taken using an observational framework and included reflexive notes.

Participant anonymity was maintained through de-identification, and data was securely stored on the National University Health System Document Repository. Access was restricted to study team members, and transcripts are archived for future research.

### Data Analysis

Data was analysed thematically using Braun and Clarke’s six-stage method, combining deductive coding based on literature findings with inductive codes derived from the data. SN, YM, AB, and LS independently conducted line-by-line coding using QSR NVivo 14. Each transcript was coded twice by two different reviewers. Disagreements were resolved by consensus or appeal to a third senior reviewer. R programming and igraph^13^ were used to visualise connections between codes, which were then categorised into themes (Appendix 5). Thematic saturation was reached before the sample size was finalised. Reflexivity was maintained throughout the study by ensuring intercoder reliability.

### Ethics Approval

Ethics approval was granted by the National University of Singapore Institutional Review Board (NUS IRB) (LS-19-143E), by Sunpasittthiprasong Hospital Ethics Committee (031/62S, CA 045/2562) and by Nepalese Health Research Council (NHRC) (359/2019).

### Role of Funding Source

The funders had no role in study design, data collection and analysis, decision to publish, or preparation of the manuscript.

## Results

### Study population and local antibiotic stewardship practices

A total of 194 interviews were conducted amongst physicians (n=105, 54·1%), nurses (n=38, 19·6%), pharmacists (n=24, 12·4%), and management staff (n=27, 13·9%). Seventy-four (38·1%) participants were from Singapore, 69 (35·6%) from Thailand, and 51 (26·3%) from Nepal (Table 1).

**Table 1.** Demographics of interview participants.

| Participant characteristics | All (n = 194) | Singapore (n = 74) | Thailand (n = 69) | Nepal (n = 51) |
| --- | --- | --- | --- | --- |
| Sex |  |  |  |  |
| Male, n (%) | 96 (49·5) | 41 (55·4) | 23 (33·3) | 32 (62·8) |
| Female, n (%) | 98 (50·5) | 33 (44·6) | 46 (66·7) | 19 (37·2) |
| Primary Role |  |  |  |  |
| Management, n (%) | 27 (13·9) | 5 (6·8) | 18 (26·1) | 4 (7·8) |
| Nurse, n (%) | 38 (19·6) | 12 (16·2) | 14 (20·3) | 12 (23·5) |
| Pharmacist, n (%) | 24 (12·4) | 10 (13·5) | 9 (13·0) | 5 (9·8) |
| Physician, n (%) | 105 (54·1) | 47 (63·5) | 28 (40·6) | 30 (58·8) |
| Years of intensive care unit experience<sup>^</sup>, median (IQR) | 5·0 (1·0 – 15·0) | 4·0 (0·5 – 10·0) | 13·0 (4·5 – 21·5) | 1·3 (0·4 – 5·5) |
| Years after graduation<sup>^</sup>, median (IQR) | 12·0 (6·0 – 19·0) | 12·0 (9·0 – 16·0) | 16 (8·0 – 28·0) | 5·0 (3·0 – 13·0) |
| Work in a Tertiary institution, n (%) | 192 (99·0) | 72 (97·3) | 69 (100·0) | 51 (100·0) |
| Prescriber, n (%) | 119 (61·3) | 50 (67·6) | 40 (58·0) | 29 (56·9) |

IQR: Interquartile range. <sup>^</sup>Sample sizes vary due to missing data: Years of intensive care unit experience - entire cohort (n=162), Singapore (n=62), Thailand (n=60), Nepal (n=40); Years after Graduation - entire cohort (n=186), Singapore (n=66).

In Singapore hospitals, antibiotic stewardship was delivered through structured programmes funded by the ministry of health where it was routine for antibiotic and antifungal prescriptions to be guided by infectious diseases (ID) specialists and pharmacists, using both front-end restrictive prescribing and back-end audits. Key performance indicators were communicated to prescribers regularly and were compared amongst departments and with other hospitals. These indicators included adherence to stewardship advice and antibiotic consumption. Some, but not all, Thai hospitals also performed front and back-end audits. The most typical model was where initial antibiotic prescriptions of broad-spectrum antimicrobials were restricted and prolonged use of certain antimicrobials required ID specialists’ approval. In Nepalese hospitals which participated in our study, there were no structured antibiotic stewardship activities during the study period. Hospitals in Nepal and Thailand were relatively understaffed compared to Singapore (Table 2).

**Table 2.** Population level data and antibiotic stewardship characteristics by country.

|  | **Singapore** | **Thailand** | **Nepal** |
| --- | --- | --- | --- |
| Country income group<sup>1</sup> | High | Upper middle | Lower middle |
| Population | 6.04 million | 71.6 million | 29.6 million |
| Physicians per 1000 population<sup>2</sup> | 2.6 | 0.9 | 0.9 |
| Hospital beds per 1000 population | 2.6 | 2.3 | 0.4 |
| Is ASP part of the National Action Plan to tackle AMR? | Yes<sup>3</sup> | Yes<sup>4</sup> | Yes<sup>5</sup> |
| Availability of ASP in the participating institutions during the study period | Yes, in all public hospitals | Yes, in some public hospitals | None |
| Is AMR programme government funded | Yes | Yes<sup>6</sup> | No |
| Antibiotics audited in the participating institutions during the study period | Most inpatient and some outpatient prescriptions | Some prescriptions, with a focus on broad-spectrum antimicrobials | None |
| Audit strategy | Both front-end and back end auditing in most institutions.<br>Front end: Prescribers required to state indication for the use of broad-spectrum antimicrobials upon prescription.<br>Back end: ID physicians and pharmacists performed personalised prescription review of broad-spectrum antimicrobial use. | Both front and back end auditing but varied by institution.<br>Front end: Request form for broad-spectrum antimicrobials required.<br>Back end: Prolonged (> 7 day) use of certain antibiotics required ID approval | None |

Abbreviations: AMR – antimicrobial resistance; ASP – antibiotic stewardship programme. Data from official sources is referenced, and remaining data in the table is extracted from interviews and ethnographies conducted in this study.

### Major themes

Across the institutions, antimicrobial prescribing behaviour was shaped predominantly by systemic factors rather than individual prescriber attributes. Three major themes emerged: (1) the importance of diagnostic, infection prevention and control (IPC), and stewardship infrastructure in supporting context-specific prescribing; (2) the influence of responsibility, hierarchy, and short-term risk on prescriber decisions; and (3) the way patient acuity intensified reliance on antibiotics when system supports were limited.

#### Theme 1. Systemic factors underpinned safe and context-specific antibiotic prescribing

Across countries, participants consistently highlighted that robust systemic support infrastructures were essential for creating a safe environment for context-appropriate prescribing decisions. Key enablers included reliable microbiology laboratories, access to high-quality antibiotics, enforcement of infection-prevention and control policies, up-to-date and locally adapted stewardship guidance, and the active involvement of allied health staff in prescribing decisions. These systemic factors interacted closely with physician related factors such as their sense of accountability towards patient outcomes, and patient related factors such as illness acuity (Figure 1).

**Figure 1.**
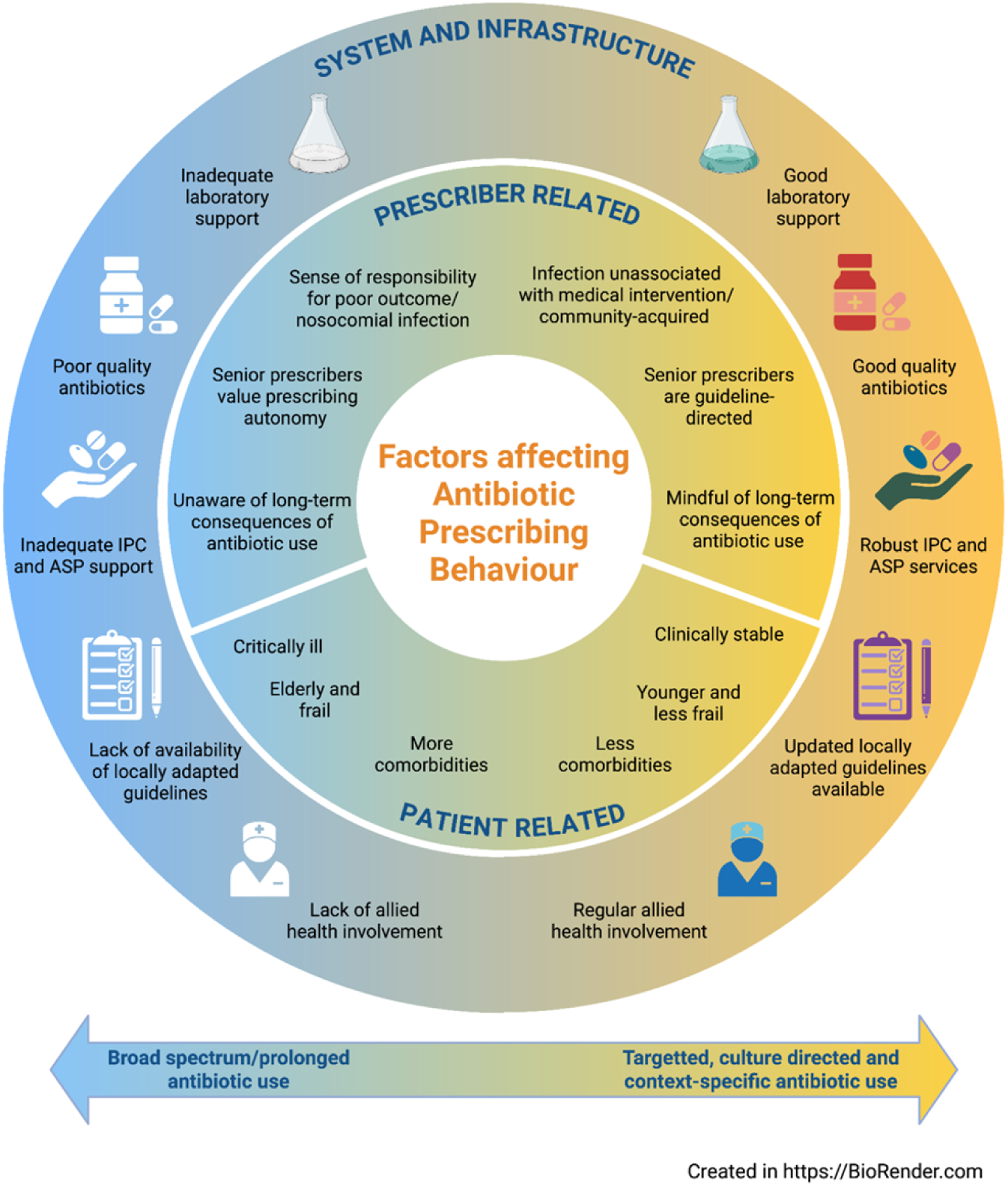
Overview of the key themes highlighting the interplay of factors between system and infrastructure (outer circle) versus individual (inner circle) determinants. Abbreviations: AMR: antimicrobial resistance; ASP: antimicrobial stewardship programme; IPC: infection prevention and control

##### 1.1: Limited laboratory reliability and antibiotic quality fuelled diagnostic uncertainty and liberal antimicrobial prescribing

Participants described how weaknesses in laboratory systems and inconsistent antibiotic quality undermined confidence in diagnostic results and treatment effectiveness. In Nepal, healthcare professionals expressed reluctance to trust negative culture results, fearing false negatives due to laboratory inadequacy (T1Q1, T1Q2). Furthermore, laboratory results often were not reported in a clinically relevant time frame and therefore did not influence antimicrobial prescribing decisions. On occasion, physicians or patients’ families had to retrieve results manually (T1Q3). Similarly in Thailand, prescribers reported that delayed or difficult-to-assess laboratory results made de-escalation of antimicrobials impractical in acutely unwell patients (T1Q4).

Concerns over antibiotic quality also shaped practice. In Thailand, mistrust in the efficacy of locally manufactured antimicrobials and weak regulatory oversight often led physicians to prescribe double doses to ensure therapeutic efficacy (T1Q5).

##### 1.2 : Weak infection prevention infrastructure heightens perceived infection risk and drives liberal antibiotic use

Participants described a close relationship between infection prevention capacity and antimicrobial decision-making. Where IPC measures were inadequate, the perceived risk of infection transmission increased, leading to earlier initiation and prolonged courses of antimicrobials.

In resource-limited institutions, ethnographic observations revealed that disposable medical equipment and cleaning solutions meant for single use were sometimes shared between patients, due to limited accessibility. Disposable, sterile items used were dependent on patients’ ability to afford these items, as they were charged for it directly (T1Q6). Non-disposable equipment such as metal scissors were dipped in cleaning solutions between each use and shared between patients. Infection prevention bundles were inconsistently implemented, and hand hygiene practices relied on shared disinfectant soaps and reusable towels. Prescribers reported that the scarcity of gowns, gloves, and other protective resources (T1Q7) increased anxiety about acquiring resistant infections. This sense of vulnerability contributed to liberal prescribing behaviours, including prophylactic or early broad-spectrum antibiotic use in high-risk patients (T1Q8). Prescribers expressed that when infection prevention measures are not in place, physicians relied on prescribing broad-spectrum antibiotics to reduce the risk of institution acquired infections (T1Q9).

In other institutions, while personal protective equipment such as gowns and gloves were generally available to healthcare workers (T1Q10), prescribers observed that lapses in IPC practices were prevalent. These lapses were attributed to infrastructural deficiencies, such as overcrowded hospital wards (T1Q11) and insufficient nursing staff dedicated solely to patient care. Such challenging conditions compromised adherence to IPC protocols, heightened the risk of pathogen transmission and also led physicians to adopt an automatic, often unrestrained, approach to antibiotic use—hesitating to discontinue antibiotics once started out of fear of patient deterioration, which exacerbated antimicrobial resistance risks.

In institutions with more stringent IPC measures, reusable equipment was autoclaved prior to reuse. IPC bundles were better established, including the use of sterile disposable equipment for every procedure performed. Sterile disposable sets, beside alcohol-based hand rubs, and single-use paper towels were readily available.

##### 1.3: Limited, inconsistent or outdated guideline support shifted reliance toward individual judgement

Across all three countries, physicians relied primarily on their own clinical judgment when making antibiotic prescribing decisions, often citing limitations in the relevance or timeliness of available guidelines. Participants noted that hospital antibiograms and institutional recommendations were infrequently updated and did not always reflect current resistance patterns (T1Q12). For critically unwell patients in particular, physicians expressed concerns that guidelines might not take into account individual patient demographics and illness acuity and were therefore insufficient in addressing specific clinical scenarios (T1Q13). Even where clear international guidelines were available, prescribers were cautious about applying them directly, as these were often developed for settings with different epidemiology and resources (T1Q14). Senior physicians, in particular, emphasised the importance of clinical experience and autonomy (T1Q15), sometimes extending antibiotic durations based on prior cases or perceived patient risk, even when such decisions diverged from standard recommendations (T1Q13). Across settings, this reliance on professional judgement reflected both reliance in individual expertise and gaps in system-level guidance to support context-appropriate prescribing.

##### 1.4 : Allied health empowerment facilitated stewardship

Across all settings, participants acknowledged the potential value of allied health professionals, particularly pharmacists and nurses, in promoting context-specific antimicrobial prescribing. However, the degree of their involvement varied depending on staffing capacity, institutional hierarchy, and stewardship infrastructure.

In Singapore, pharmacists and nurses were more consistently felt empowered to contribute to prescribing decisions (T2Q16). Though variable, they participated actively in the ward rounds by conveying the patients’ clinical condition to the physicians or enquiring about the planned antibiotic duration. Pharmacists in Singapore had some autonomy, with dedicated antimicrobial stewardship pharmacists who reviewed patients records and provided tailored guidance for inappropriate antibiotic prescriptions on medical records with support from ID specialists.

In Thailand and Nepal, pharmacists verified and dispensed antibiotics, and advised on appropriate dosing and administration (T1Q17, T1Q18). Their participation in daily rounds or prescribing discussions was limited, often due to heavy workloads or traditional role delineations. Pharmacists in Nepal were especially short staffed, with one pharmacist checking manual prescriptions for multiple wards. Nurses likewise contributed to infection monitoring and antibiotic administration, though hierarchical team structures sometimes constrained their ability to influence prescribing decisions (T2Q19, 20, 21).

#### Theme 2. Individual prescribing agency constrained by immediate clinical imperatives and hierarchical culture

While physicians’ sense of accountability to their patients’ immediate well-being and professional culture strongly influenced antibiotic decisions, these behaviours were often shaped by system-level norms and organisational hierarchies. Prescribers operated within environments that rewarded short-term risk aversion and deference to authority, making it difficult to consistently align practice with stewardship goals.

##### 2.1 Perceived responsibility towards patient and infection shaped prescribing focus

Physicians’ sense of responsibility was closely tied to the source and perceived ownership of an infection. Surgeons, for example, described heightened accountability for post-operative infections, prompting them to initiate antibiotics proactively to protect surgical outcomes (T2Q1). Conversely, for conditions viewed as outside of surgeons’ domain, such as ventilator-associated pneumonia, surgeons were more willing to defer to ICU or ID specialists (T2Q2), recognising that these professionals were specifically trained to navigate such infections and ongoing treatment considerations.

This sense of ownership sometimes led physicians to deviate from stewardship recommendations or multidisciplinary advice, motivated by the fear of adverse outcomes. Primary physician teams perceived themselves as most accountable for patient survival and were hesitant to accept interventions from other clinicians or pharmacists (T2Q3). In Singapore, although there was a more structured approach to antibiotic stewardship, some physicians resisted antibiotic review recommendations, citing personal responsibility for treatment failure as justification for overriding external input, particularly in the ICU setting (T2Q4).

##### 2.2 Seniority and prescribing culture overrode guideline-based practice

Across all sites, senior clinicians exerted substantial influence over antibiotic decisions, setting the tone for team prescribing culture. Junior physicians often deferred to senior consultants’ preferences, viewing them as more experienced and better equipped to judge risk (T2Q5, Q6, Q7). This hierarchical dynamic sometimes displaced evidence-based guidelines, with compliance to authority taking precedence over adherence to protocols (T2Q8). The fear of repercussions for not following senior directives drove many to adhere to potentially suboptimal prescribing patterns.

##### 2.3 Immediate benefits of antibiotics outweighed perceived long-term risks of resistance

Physicians commonly prioritised the tangible, short-term benefits of antibiotic therapy, such as rapid clinical stabilisation and prevention of deterioration, over the abstract, long-term risks of antimicrobial resistance (T2Q9). Some participants expressed concern about the repercussions of AMR, sharing personal experiences of managing patients who suffered poor outcomes due to resistant infections (T2Q10). Others felt that the true impact of AMR was not yet perceptible in their day-to-day practice (T2Q11). The risks associated with AMR at the population level and in the future were viewed as diffuse and probabilistic, seldom directly linked to individual prescribing decisions, much like climate change (T2Q12). This perceived disparity between immediate benefits and potential harm fostered a tendency to favour initiating broad-spectrum or prolonged antimicrobial therapy, especially in high-acuity settings.

#### Theme 3: Patient-related factors influencing antibiotic prescribing

##### 3.1: Patient demographics and illness acuity created high-stakes contexts that strongly influenced antibiotic prescribing decisions

Prescribers often resorted to longer antibiotic durations in the elderly or frail, those with more co-morbidities (T1Q17), or the critically ill, without clear evidence of the benefit of this approach. In the ICU setting, the immediate clinical priority for physicians was to stabilise critically ill patients and facilitate their transfer to the general ward. The high stakes of patient care led physicians to prioritise empirical antibiotic treatment (T3Q1), as delays could result in adverse outcomes (T3Q2). Consequently, ICU practitioners used broad-spectrum antibiotics to cover all potential pathogens, given the unpredictable evolution of infections in critically ill patients. Physicians also tended to have a higher threshold to stop antibiotics in the ICU setting, where the consequences of premature cessation of antibiotics could be dire (T3Q3). Such practices may be prevalent even if a focus of infection was unclear (T3Q1) as patients in the ICU setting can have undifferentiated syndromes.

**Table 3.** Quotes during interviews of healthcare staff and ethnographies.

| Theme number, quote number | Quote | Role, years since graduation, country |
| --- | --- | --- |
| T1Q1 | "I don't feel that much confidence (in de-escalating antibiotics) because I don't feel that our lab is strong. When we send the sputum which is yellow or greenish sputum for which we are sure there will be growth, but there is no growth, I feel our lab is not strong." | Junior physician, 5 years, Nepal |
| T1Q2 | "If the patient is improving clinically then we have continued (antibiotic to which pathogen is resistant), because we can't just focus on the lab factor. We have to mainly look at the patient's clinical condition. We don't know how that happened, if it was a lab error or not, but it has happened" | Junior physician, 3 years, Nepal |
| T1Q3 | A (Consultant) asked for the report of cerebrospinal fluid which was sent the night before. J (medical officer) answered that the report was not collected. A seemed disappointed and said "if you are doing any invasive procedure, the report should be with you within an hour otherwise you have no right to do the procedure during off hours (night)." A also mentioned "visitors are not responsible for patients, you are" after the Medical Officer in ICU said that they have sent visitors to collect the report. | Ethnography during morning medical conference, Nepal |
| T1Q4 | "Maybe after five days we still don't know the culture results. So, we can't de-escalate." | Manager and Physician, 29 years, Thailand |
| T1Q5 | “In our country, the first problem is that there is no standardised drug. Because the drugs that we use are locally made drugs, in which their companies are not standardised, but we must give antibiotics – it's like we have to close one eye – because our budget is small. We solve the problems by giving a double dose of antibiotics.” | Manager and Physician, 30 years, Thailand |
| T1Q6 | A (Consultant) asked “how much for the dressing set”? Nobody answered it and he said “it's around 200 or 300 Rupees only why we are not using sterile dressing set all the time?” | Ethnography during MICU rounds, Nepal |
| T1Q7 | “It's (PPE) a bit insufficient. There are instances when it's not enough since there are lots of people during the rounds, and sometimes the gown used in the morning is reused again and again, which in itself is a source of infection. They (doctors) wear the same gown and go from patient to patient or someone else wears the same gown and goes to another patient.” | Junior physician, 3 years, Nepal |
| T1Q8 | “If they would provide these (gloves, masks, gowns) it would be good but it would be difficult for the patient to bear the expenses, because they are usually from economically poor backgrounds. Because of such conditions we (our patients) are at high risk of infections with antimicrobial resistant pathogens...(to overcome this) sometimes in the ICU we do not leave patients on a ventilator without prophylactic antibiotics.” | Junior physician, 1 year, Nepal |
| T1Q9 | “If infection control is good with good infrastructure then antibiotics will not be prescribed. Only ceftriaxone will be given even in brain surgery. Otherwise, if infection control is not done well then patients will be prescribed clindamycin, flucloxacillin along with ceftriaxone.” | Senior physician, Anaesthesia, 10 years, Nepal |
| T1Q10 | As an infection prevention and control nurse, I focus on the implementation of measures for prevention of antibiotic-resistant organisms. The first thing that I emphasise is washing hands. Second is isolating them (patients with resistant organisms) into separate rooms. Then, we make arrangements for the equipment, which will be worn in caring for the group of infected patients, namely waterproof coats and gloves. We will separate equipment, such as stethoscopes, BP cuffs, baskets, bedside cabinets where items are kept and used by the patient.” | Nurse, 15 years, Thailand |
| T1Q11 | “The authorities need to take the lead and say - I think the ICU beds can't be 12 beds; it should have only 6 beds, and each bed should be taken care of by two nurses. If we can monitor and keep standards like this, the problems would be solved. But currently it is a slum with air-conditioning and patients are crowded like canned fish. For example, when performing suctioning (in ICU), the pathogen goes across four beds, which are cramped together. This way, you introduce an automatic antibiotic use system. No one dares to stop the antibiotics. Really, do you dare if you are an interventionist? I performed surgery for 2- 3 times, and then the patient died due to stopping antibiotics. So, I don't want to take a risk. So, we really have to improve our standards.” | Senior physician, Surgery, 29 years, Thailand |
| T1Q12 | “Regarding the information used as a guideline, it is not up to date. Even the antibiogram, we update it very slowly. We wait until the end of the year to obtain the results of that year.” | Manager and physician, 29 years, Thailand |
| T1Q13 | “Yes, even now as I am going through my higher training there are differences in how consultants prescribe antibiotics, some give 5 days for a similar infection as compared to someone that would give a duration of 2 weeks and there really is no good evidence for a longer duration. And it is sort of just based on how the guy has been trained and, and I guess in some ways we might try to justify it by saying, this patient is older, a bit more co-morbidities and it's probably safer to give it for a longer duration whether or not that is evidence based.” | Junior physician, HPB surgery, 9 years, Singapore |
| T1Q14 | When asked whether they would adopt research findings from other countries – “Oh, this is even more difficult. There will certainly be different microbiome.” | Junior physician, 3 years, Thailand |
| T1Q15 | When asked if their clinical practise would be influenced if a randomised controlled trial found no difference in long or short antibiotics duration in patients with HAP – “There will be no change in service behaviour for sure. No change. My medical profession license, it's an art – the art that has been taught this way. Yours may be the first research paper or one of the most popular papers, but it only lasts for a short duration. But in the next ten years, another generation of doctors can change and implement this.” | Senior physician, Infectious diseases, 20 years, Thailand |
| T1Q16 | “We do the rounds with doctors; it is more like a collaborative approach. I would say we proactively look out for the usage of antibiotics, making sure that it is not started without any reasons. And if it's started we check if there's an end date.” | Nurse, 19 years, Singapore, |
| T1Q17 | When asked if pharmacist input would be helpful in antibiotic prescribing – “There is no difference because the judgement whether to continue or discontinue the drug still belongs to the doctor. They (pharmacists) can help with antibiotic dosing.” | Junior physician, 4 years, Thailand |
| T1Q18 | When asked if a multidisciplinary team (pharmacists) should be involved in antibiotic prescribing and how so – “First, they are to determine whether the dose is appropriate or not. Second, (they can guide on) solvents that are used to mix antibiotics. Sometimes we forget how the antibiotics are supposed to be mixed.” | Junior physician, 3 years, Thailand |
| T1Q19 | “As a nurse, we’re quite involved in giving advice to the doctors, for instance, how the sputum is and how its colour is now. We provide information for them to consider. But we cannot tell them to stop right away – everyone has their own ego.” | Nurse, 20 years, Thailand |
| T1Q20 | “For the nurses, they’re involved in this (antibiotic prescribing) very little; their focus is on correctly managing according to the doctor’s orders preventing a patient from contracting an antibiotic-resistant organism from the neighbouring contagious case” | Nurse, 33 years, Thailand |
| T1Q21 | When asked if they (nurses) influenced antibiotic prescribing – “No we do not” | Nurse, 7 years, Nepal |
| T2Q1 | “Most of the surgeons when I tell them to stop anything they are not willing to stop. I do understand their anxiety because they are the ones who did the surgery and if anything happens it comes back to them you see” | Senior physician, Infectious diseases, 12 years, Singapore |
| T2Q2 | “I suppose in a Neurology ICU patient, the surgeons are more concerned about patients that have post-surgical infections like with extra-ventricular devices or shunts or recent craniotomies. For surgical patients the surgeons may not really even be aware when we suspect ventilator acquired pneumonia because it would not be something that is part of their surgical management. So that would be under the ICU and we would have a bit more responsibility whether or not we escalate antibiotics rather than the surgeons asking to escalate antibiotics” | Senior physician, Anaesthesia, 10 years, Singapore |
| T2Q3 | “They (ASP) come and ask us to stop that, isn’t it? That’s all based in their judgment, right? That should actually be done under the judgment of those who have been looking after patient” | Senior physician, 9 years, Nepal |
| T2Q4 | “You would have to be prepared for a lower-than-expected (ASP) acceptance rates from the physicians, if you’re talking about the ICU where there are higher stakes involved in patient care.” | Senior physician, Infectious diseases, 11 years, Singapore |
| T2Q5 | “Due to the fear of full responsibility of patient’s life on their shoulders consultants do not follow guidelines as well as residents do” | Junior physician, 4 years, Nepal |
| T2Q6 | “Residents and MO’s usually follow the guidelines. However, some consultants don’t follow it. Maybe they haven’t read the guidelines or due to the fear of full responsibility of patient’s life in their shoulder the consultant does not follow guidelines as residents do.” | Junior physician, 5 years, Nepal |
| T2Q7 | “I worked at a hospital where there are more cases of myocardial infarction (MI). Patients with MI definitely have leucocytosis but they start antibiotics for presumed infection even if the count is mildly elevated. I couldn’t challenge and speak there; I wasn’t comfortable to suggest anything due to the presence of consultants and professors.” | Junior physician, 3 years, Nepal |
| T2Q8 | “That’s the kind of practice culture around here. Everyone just does what the consultants want. Let’s say I’m the registrar on call, I will be handing over the case that I saw overnight to the consultant the next morning. I could definitely foresee myself getting into trouble for not starting an antibiotic but it would be almost unthinkable that they would call me to question why I gave an antibiotic.” | Senior physician, Internal medicine, 9 years, Singapore |
| T2Q9 | “The honest truth is that I think people know it (about AMR). But people will err on the side of caution because you do not want them to deteriorate. The outcomes and consequences of antimicrobial resistance, plays a small factor in the person's clinical decision making process when the patient is critically ill.” | Senior physician, ICU, 12 years, Singapore |
| T2Q10 | “I’m worried, I’m extremely worried, because this (antimicrobial resistance) has affected the field I’m in. I’ve seen people die because of multi-drug resistant organisms. I’ve seen its effect; its economic burden, its social burden, and because of all those concerns it’s a matter worth worrying about.” | Senior physician, Professor of microbiology, head of infection control, 10 years, Nepal |
| T2Q11 | “When we talk about resistance it's not a problem that is very palpable. Meaning that a lot of us won't see it. And a lot of us will not experience the difficulty of dealing with a patient with resistance so that's I think one of the main reasons why a lot of us don't think about it. I guess it's human nature if we don't see the true consequence it's quite hard to convince us to do it.” | Senior physician, Colorectal surgery, 9 years, Singapore |
| T2Q12 | “Right now I’m healthy, I’m not immunocompromised, so even if I have MRSA or VRE it does not affect me, it doesn’t affect my day-to-day life, it doesn’t affect my family or my friends right now even though I know logically intellectually if somebody else is sick and they get it from me, then that would be devastating for them. It’s like climate change - you know it’s very important, but it’s difficult to motivate yourself at this point in time. | Senior physician, Anaesthesia, 10 years, Singapore |
| T3Q1 | “Usually in our ICU all the patients that we get are critical patients, so we usually use broad antibiotic coverage, even if there is no culture growth, or focus of infection. In the case where we don’t know which bacteria is causing the pneumonia, we take all the prevailing bacteria in Nepal into consideration....like from *Staphylococcus aureus* to atypical bacteria, we try to cover them all” | Junior physician, 1 year, Nepal |
| T3Q2 | “In VAP cases, if we delay treating them, we will surely kiss them goodbye.” | Junior physician, 8 years, Thailand |
| T3Q3 | “If the patient is still clinically critically ill my threshold to want to stop antibiotics would be very high.” | Senior physician, ICU, 14 years, Singapore |

Senior physicians are defined as those who have obtained specialist accreditation. Junior physicians consisted of those who were not in specialist training or have yet to complete specialist training at the time of their interviews.

## Discussion

In this multinational qualitative study across healthcare settings of various income levels, we found that antibiotic prescribing in institutions was shaped primarily by system-level support, including the robustness of IPC, diagnostic capacity and antibiotic quality. Where these infrastructures were strong, they created a safer clinical environment that enabled more targeted prescribing by reducing individual clinician’s fear of patient deterioration. In contrast, in settings with weaker system support, prescribing decisions were strongly driven by patient acuity and prescriber risk aversion, resulting in a tendency toward cautious and prolonged antibiotic use. These findings suggest that prescribing behaviours often labelled as “inappropriate” or “defensive” may instead represent rational responses to uncertainty within the health system, rather than deficits in knowledge or motivation.

The role of infrastructural and support services in enabling context-specific antibiotic prescribing has received less attention than individual or interpersonal influences in published literature. This imbalance likely reflects the fact that much of the qualitative literature on prescribing behaviour originated from high-income settings, where IPC, microbiology diagnostics, prescribing guidelines, and antibiotic quality were largely taken for granted^11,14,15^. However, in resource limited settings, access to timely and reliable microbiology support is often limited^16, 17, 18^. Delays or uncertainty in laboratory reporting constrain clinicians’ ability to assess patients’ clinical trajectories and adjust therapy, especially in critically ill populations. This is further compounded by frequent lack of availability of high-quality drugs due to unreliable supply chains and poor regulatory oversight^19^. In the absence of comprehensive, locally relevant guidelines, clinicians frequently rely on personal experience to guide prescribing decisions. Furthermore, poorly established IPC measures contribute to spread of AMR which may make physicians more fearful of poor patient outcomes due to serious infective complications. This then drives broad-spectrum antibiotic prescribing^20^. By shifting attention from individual decision-making to the conditions under which decisions are made, our study extends existing literature that has predominantly focused on prescriber knowledge, attitudes, and interpersonal dynamics. This also highlights how responsibility and risk are distributed across individuals and institutions, further reinforcing the need to consider stewardship as a shared, system-level function rather than an individual responsibility.

Another important insight from this study was that while prescribers were fundamentally driven by a deep-seated desire to achieve the best possible outcomes for their patients, responsibility for antibiotic decision-making shifted depending on who was perceived to “own” the infection. For example, surgeons could tend to approach surgical site infections with a focus on protecting operative outcomes, whereas infectious diseases specialists were more attuned to treatment duration, downstream resistance, and broader patient and public health implications. These differing priorities reflect the distinct roles clinicians occupy within multidisciplinary teams and illustrate how professional identity and perceived accountability shape prescribing behaviour in context-specific ways. This finding complements prior work by Charani *et al*, which highlighted substantial heterogeneity amongst prescribers and the need for more targeted interventions amongst physicians of different specialties with different agendas, rather than applying uniform interventions irrespective of clinical role or agenda^21^.

Our findings suggest that effective antimicrobial stewardship should be reframed as a problem of system design rather than individual behaviour correction, especially in resource limited settings. Current interventions described to improve antimicrobial prescribing in the institution settings have largely focused on modifying individual prescriber behaviour through education, audits and feedback, and drug-restriction policies^22–25^ as well as resource intensive strategies such as rapid gene-based diagnostics and personalised antimicrobial stewardship services^26^. While these approaches may be effective in well-resourced settings, they often fail to address the systemic constraints faced by low- and middle-income countries and can be difficult to implement or sustain in resource-limited environments. Taken together with existing evidence, our findings support a shift toward strengthening foundational health-system capacities, including diagnostics, IPC, antibiotic supply, and stewardship governance, as key enablers of appropriate prescribing. To make a meaningful impact, the focus needs to shift to enable these settings to strengthen laboratory support including laboratory management systems, improve the availability of good quality drugs, establish basic IPC guidelines and practices, and improve accessibility to updated and locally relevant guidelines. Such measures are critical to improve physician trust in the healthcare system and reduce the burden on individual prescribers in making antibiotic prescription decisions. This system-oriented approach may be particularly important for achieving sustainable improvements in antibiotic use in low- and middle-income settings, where behaviour-focused interventions alone are unlikely to succeed.

The major strength of this study was that it brought together perspectives from different professional groups across countries of varying income levels, using interviews alongside ethnographic observations, which provided triangulation of findings and improved the credibility of the analysis. We also acknowledge the limitations. Data were collected from selected institutions and clinical areas, and therefore might not have captured the full range of prescribing contexts within each country. In addition, while the study focused primarily on hospital and ICU settings, prescribing dynamics might differ in other care environments. Finally, although the cross-country design strengthens conceptual transferability, local organisational cultures and policies may limit direct generalisation of specific observations.

## Conclusion

Our findings suggest that behaviour change interventions for rationalising antibiotic prescribing should extend beyond individual behaviour change and focus on building system conditions that allow clinicians to prescribe safely. Creating a supportive infrastructure through reliable diagnostics, effective infection prevention and control, assured antibiotic quality, and embedded stewardship, reduces uncertainty and shifts responsibility from individual prescribers to the health system. Such a holistic approach is essential for enabling rational, context-specific antibiotic use.

## Data Availability

Interview transcripts will be available after acceptance.

## Declaration of interests

We declare no competing interests.

## Acknowledgement

The project was funded by Medical Research Council/ Department for International Development (MRC/DfID) (Grant Ref: MR/K006924/1) and Singapore National Medical Research Council (Grant Ref: CTGIIT18MAY-005)

## Authors and Contributors

SC, SN, YM, CJY, DRS analysed the data and wrote the manuscript. AYL, GK, PC, YM recruited participants for this study. AB, KV, CJY performed the interviews for this project. LS, YM, AB, SN and AHC independently performed coding for each transcript and ethnography. BSC, EC reviewed the manuscript. YM supervised the entire project.

## Data Sharing

Data collected for the study, including de-identified participant data, data dictionary, and additional related documents, will be made available to others upon request to, following the Mahidol-Oxford Research Unit’s data sharing policy and in accordance with WHO statement on public disclosure of clinical trial results.

## Notes

### Competing Interest Statement

The authors have declared no competing interest.

### Clinical Trial

NA

### Funding Statement

Yes

